# Association of Cognitive Abilities with Patterns and Quality of Sleep in Nurses Working in Public and Private Tertiary Care Hospitals: A Cross-sectional Study in Pakistan

**DOI:** 10.1101/2025.05.02.25326876

**Authors:** Anusha Azhar, Muhammad Maaz Bin Zahid, Sunnia Shah, Rida Islam, Sakshi Kumari, Muhammad Sanaan Noor, Aina Khushbakht Tariq, Bushra Iftikhar

**Affiliations:** Department of Medicine, Khyber Medical College, Peshawar, Pakistan; Chairperson, Department of Community Medicine, Khyber Medical College, Peshawar, Pakistan

**Keywords:** Sleep quality, sleep patterns, cognition, health-care workers, nurses

## Abstract

Healthcare workers, especially the nurses are one of the most overworked individuals of the society. Huge working hours can affect the quality and patterns of sleep, which in-turn can affect the cognitive abilities of these workers. The primary aim of this study was to determine and evaluate the impact of sleep quality and sleep patterns on cognitive abilities of nurses in tertiary care hospitals. The secondary objective was to ascertain differences in cognition and sleep quality of different shift nurses of public and private tertiary care hospitals. This cross-sectional study was conducted on 360 nurses across six tertiary care hospitals in Peshawar, Pakistan. Sleep quality was assessed using the Sleep Quality Scale (SQS) scale. Sleep patterns were analyzed using a self-administered questionnaire. Cognitive abilities were quantified using the Mini-Mental State Examination (MMSE). Statistical tests included were the Mann-Whitney U, Kruskal Wallis, and Spearman’s correlation test. Out of the 360 nurses, 54.2% of them were females, with a median age of 27.7 +/- 5.6 years. The median SQS and MMSE were 41 ± 91, and 23 ± 30. Significant differences between public and private hospitals were noted in work hours, night sleep time, and nap duration (p<.05). For shift nurses, it included work hours, and MMSE (p<.05). Sleep pattern factors associated were nap duration (ρ =0.12), and frequency of sleep cycle (ρ =-0.24). Moreover, there was a negative correlation between SQS and MMSE (ρ =-0.024). This study concludes that sleep quality directly influences the cognitive functioning of nurses. Among the sleep patterns, nap duration and frequency of sleep cycle were associated with cognitive abilities. Additionally, significant differences in the distribution of work hours, night sleep time, and day shift frequency were observed.

## 1.0 Introduction

Sleep is a naturally occurring essential process characterized by altered levels of consciousness and decreased bodily movement and responsiveness to external stimuli.[1] It is an essential component that plays a pivotal role in the physical, mental, and emotional well-being in the course of human life. It is a complex and dynamic process that involves a multitude of physiological and neurological mechanisms, ultimately facilitating rest and regeneration. Sleep is essential for many bodily functions, including the immune system, metabolism, and cardiovascular health. An average human spends about one-third of their life sleeping.

In addition to its restorative and metabolic functions, sleep has a close link to a range of cognitive processes, including memory consolidation, learning, and decision-making, depending on the specific conditions of learning and the timing of sleep. [2] Several studies have explored the association between sleep patterns and cognitive abilities. Research conducted on a group of adolescents proved that napping can enhance declarative memory.[3] Another study highlighted that loss of sleep can have a significant impact on cognitive functioning.[4] Much attention has been paid to the role of sleep in decision-making,[5] innovative thinking,[6] alertness, and execution of simple tasks, where all investigations consistently indicate enhanced performance in these domains in the company of optimal sleep.[7].

Globally, the prevalence of cognitive impairment ranges from 16 to 22.2%, with most of the patients being the older population.[8] One study observed a 40.0% prevalence of cognitive impairment in males and 45.1% in females. Furthermore, a significantly higher prevalence after the age of 75 has been reported in females.[9] Age-related changes in sleep have been extensively studied, and the correlation between these changes and the effects on cognitive decline, both normal and pathological, has been explored in research.[10]

Not just the quantity and quality but the timing of sleep also plays a crucial role in cognitive functioning. A study conducted in Iran found that, after adjusting for age and sex, one of the factors that had a significant impact on daytime functioning was the sleeping time, and duration of sleep. The study also indicated a significant rise in the prevalence of sleep disorders in recent years, with 9 to 15% of adults suffering from chronic sleep disturbances reporting daytime impairments.[11] A systematic review on napping in older adults observed that the frequency of daily naps increases with age, with the prevalence ranging from 20% to 60% worldwide.[12] Furthermore, a study associated sleep apnea with cognitive decline, indicating that decreased sleep quality may become a risk factor for several psychiatric illnesses and can potentially aggravate cardiovascular problems.[13]

Extensive research has been conducted on how sleep patterns can affect cognition in healthcare professionals. A systematic review on the impact of sleep and circadian disorders on physician burnout observed that sleep insufficiency and exhaustion are associated and prevalent in healthcare workers, including nurses, trainees, and practicing physicians.[14] A study on sleep quality and its relationship with cognitive abilities among 540 nurses (66.3% female) working in six hospitals concluded that poor sleep quality can negatively impact both patients and nurses. It highlighted that 77.4% of the sample population had poor sleep quality, with evidence suggesting that enhanced cognitive processes can improve the sleep quality of these nurses.[15] A survey of insufficient sleep and its effects on cognitive abilities of residents and attending physicians was conducted in Jerusalem in 2020, which suggested that severe insomnolence was associated with decreased cognitive processing.[16]

The available global literature reinforces the association between sleep and cognition in nurses. However, data from Pakistan and specifically from the province of Khyber Pakhtunkhwa (KP), which has its own peculiar socio-cultural and ethnic characteristics, remains limited. This study is significantly relevant to Pakistan’s population, mirroring global trends, and is expanding at a faster pace than the number of available healthcare workers. Consequently, all healthcare professionals – especially nurses, who form the backbone of hospital care - are overworked. Erratic shift patterns, extended working hours, and an upward trend of assigned shifts contribute to nurses’ declining sleep quality, leading to poor work performance. Keeping both the immediate and long-term implications of this scenario in mind, the primary objective of this research was to determine if sleep quality and sleep patterns have any impact on the cognitive abilities of nurses in tertiary care hospitals. The secondary objective was to ascertain differences in cognition and sleep quality of different shift nurses and amongst those employed in public and private tertiary care hospitals.

## 2.0 Methodology

### 2.1 Study design and setting

This cross-sectional study was conducted from February to July 2024 across six tertiary care hospitals in Peshawar, Pakistan: Khyber Teaching Hospital (KTH), Hayatabad Medical Complex (HMC), Lady Reading Hospital (LRH), Rehman Medical Institute (RMI), Peshawar Institute of Medical Sciences (PIMS), and Northwest General Hospital (NWGH). The study followed the Strengthening the Reporting of Observational Studies in Epidemiology (STROBE) guidelines.

### 2.2 Study population and sample

The study population consisted of healthy staff and internee nurses, between the ages of 20 to 59 years, working in public and private tertiary care hospitals of Peshawar. Informed consent was taken from the nurses for their participation in the study. Nurses between the ages of 20 to 59 years old who consented to participate in the study were included. Participants with neurologic (such as insomnia, narcolepsy, restless leg syndrome, and sleep apnea) or psychiatric issues, a history of medication affecting sleep or cognition, terminal/chronic illnesses, or drug abuse were excluded.

#### 2.2.1 Sample size

The sample size was calculated using the formula of single proportion, which is as follows:

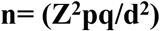

Here, n is sample size, Z (standard normal variate), is 1.96 for a 95% confidence interval, p (expected prevalence/ proportion), is 62% (0.62), calculated by taking the average prevalence of sleep disorders among nurses from multiple regional studies, [17,18] q= 1-p, and d (level of error), is taken as 5%. After applying the formula and adding a 10% non-response rate, a sample size of 356 participants is determined. A total of 360 nurses were recruited for the study. A selection flow chart of participants is given in **Fig. 1**.

**Fig 1:**
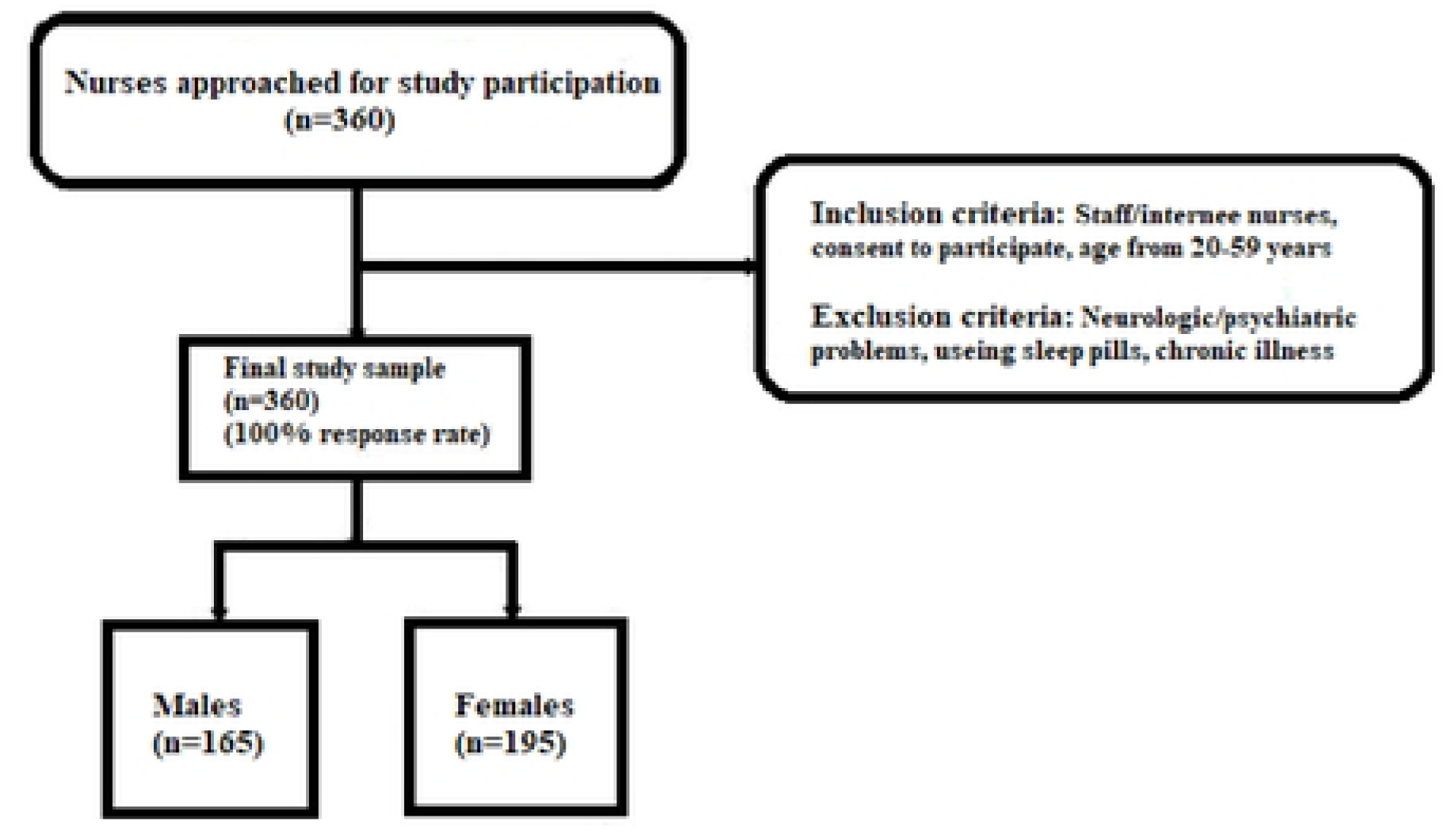
Flow chart of the study.

#### 2.2.2 Sampling method

A multistage sampling method was implemented for the selection of the representative sample. Initially, simple random sampling was employed from a sampling frame comprising all the public and private tertiary care hospitals located in Northern, Central, and Southern Peshawar, Khyber Pakhtunkhwa (KP). Finally, three public (KTH, HMC, and LRH) and three private hospitals (RMI, NWGH, and PIMS) were chosen. The selected hospitals are the primary healthcare providers in Peshawar with an extensive incoming patient volume.

Consequently, twenty-four wards, four from each hospital, were randomly selected. A list of nurses from each hospital was obtained from the information and technology department. As the distribution of nurses in each of these hospitals was nearly equal, for a uniform representation, 60 nurses were recruited from each hospital. To attain the required sample size of 360, participants were selected from several hospital departments (surgical, pediatrics, medical, gynecology, cardiology, neurology) based on inclusion criteria. The sample distribution method for each hospital is elucidated in **Supplementary file 1**.

### 2.3 Data collection tools

#### 2.3.1 Assessment of Sleep Quality

The sleep quality of nurses was assessed using the Sleep Quality Scale (SQS) **(Supplementary file 2, P1)**, a validated tool[19] developed by Hyeryeon Yi, having a Cronbach’s alpha coefficient of 0.92 for internal consistency. The SQS scale is composed of 28 items related to the quality of sleep, with each item having four options. It is a Likert scale composed of 4 points. The range of scores is from 0 to 84. Higher scores indicate lower sleep quality.[19]

#### 2.3.2 Assessment of Sleep Patterns

Sleeping patterns were evaluated using a self-administered scale **(Supplementary file 2, P2)**. A pilot study was conducted under established protocols to strengthen the reliability of the following scale components: night sleep time, waking time, night sleep duration, nap duration, and frequency of sleep cycle. The study was conducted under the supervision of a trained researcher, the results were thoroughly discussed, and the data was scrutinized by a trained data analyst, which produced satisfactory results. All the components of the sleep pattern questionnaire were further subdivided into ordered sections to assess the sleep patterns of nurses.

#### 2.3.3 Assessment of Cognition

Cognitive functioning of nurses was assessed using the Mini-Mental State Examination (MMSE) **(Supplementary file 2, P3)**, a validated tool developed by M. F. Folstein et al.[20] The MMSE score ranges from 0-30 with scores greater 25 interpreted as normal cognitive status. A lower score indicates poor cognition and intellectual skills. The scale demonstrates a sensitivity of 81% (95% CI, 78% to 84%) and a specificity of 89% (95% CI, 87% to 91%).[21.

### 2.4 Patient and public involvement

Patients and/or the public were not involved in the design, conduct, reporting, or dissemination plans of this research.

### 2.5 Statistical analysis

The collected data was analyzed and managed using the Statistical Package for the Social Sciences (IBM SPSS V.22.0). Descriptive statistics were presented as frequencies and percentages for categorical variables, and median ± interquartile range (IQR) for continuous variables. The Shapiro-Wilk test confirmed the distribution of continuous variables to be non-normal, accordingly, non-parametric tests were applied for advance analysis.

The association between the categorical variables and hospital type as well as shifts, was established using chi-square test. Non-parametric tests, like the Mann-Whitney U test, were employed to associate continuous variables with hospital type, while Kruskall-Wallis test was used to relate continuous variables with shifts.

The correlation between sleep patterns and cognition, as well as sleep quality and cognition was assessed using Spearman’s correlation test. A scatter plot demonstrates the results of the association between sleep quality and the cognition of nurses. A probability (p) of <0.05 and a CI of 95% were considered for statistical significance.

### 2.6 Ethical Issue

All participants provided informed consent and were fully guided about the study protocols and purpose. Ethical approval was obtained from the Institutional Research and Ethical Review Board (IREB) of Khyber Medical College (KMC) on 13^th^ February 2024 (No. 87/DME/KMC). The research was conducted while observing ethical guidelines, and a structured questionnaire was employed during the study.

## 3.0 Results

### 3.1 Participant characteristics

Our results show that out of the 360 nurses, 195 (54.2%) of them were females. The median age of the participants was 26±31 years. Staff nurses were dominant in prevalence, with 304 (84.4%) nurses. The median SQS value of the participants was 41± 91, while that of MMSE was 23± 30. The results of the Chi-square analysis show that the following factors between public and private hospitals were significant: work hours, night sleep time, waking time, and nap duration. The majority of the public hospital nurses on average were working for less than 50 hours per week (80%) compared to private hospital nurses (59%) (p<0.001). Private hospital nurses on average were sleeping before midnight (83.3%) compared to public hospital nurses (66.7%) (p<0.001). Private hospital nurses on average were also waking up before 8 am (94.4%) in comparison to public hospital nurses (81.7%) (p<0.001). Private hospital nurses were much more likely to partake in daytime naps (77.8%) in contrast to public hospital nurses (53.3%) (p<0.001). The Chi-square analysis showed that the results of both work hours and night sleep time amongst shift workers (morning, evening, night) were significant. The higher number of evening shift nurses were working on average for less than 50 hours per week (77.6%) compared to a morning shift (73.9%) and night shift nurses (40.9%) (p<0.001). Both morning shift (77.6%) and evening shift nurses (78.8%) on average were sleeping before midnight compared to night shift nurses (54.5%) (p=0.02). The results of the Mann-Whitney U test show that the factors having differences in hospital distributions were day shift frequency (median±IQR) (p=0.01), and night shift frequency (median±IQR). Private hospital nurses on average had lesser day shift (16±30) and night shift frequency (10±30) compared to public hospital nurses (day shift= 20±30, night shift frequency= 10±21) (p<0.001). The results of the Kruskall-Wallis test show that the factors that proved to be significant in shift nurses were, age (p=0.02), night shift frequency (p=0.02), and MMSE scores. The evening shift nurses had the highest MMSE scores (median ±IQR) (24±30), followed by morning shift (23±21) and night shift nurses (20±15) (p<0.001). The values are arranged in **Table 1**.

**TABLE 1:** Baseline and Clinical Characteristics.

|  |  |  | n (%) | Hospital |  |  | Shift |  |  |  |
| --- | --- | --- | --- | --- | --- | --- | --- | --- | --- | --- |
|  |  |  |  | public (n) | private (n) | p | morning | evening | night | p |
| 1 | Gender | female | 195 (54.2%) | 101 | 94 | 0.45 | 139 | 36 | 20 | 0.45 |
|  |  | male | 165 (45.8%) | 79 | 86 |  | 111 | 30 | 24 |  |
|  |  |  |  | public<br>(n) | private<br>(n) | *p* | morning | evening | night | *p* |
| 2 | Post | staff | 304<br>(84.4%) | 154 | 150 | 0.47 | 204 | 61 | 39 | 0.24 |
|  |  | internee | 55<br>(15.3%) | 25 | 30 |  | 45 | 5 | 5 |  |
| 3 | Work hours | <40 | 51<br>(14.2%) | 34 | 17 | <0.001 | 38 | 8 | 5 | <0.001 |
|  |  | 40 - <50 | 203<br>(56.4%) | 113 | 90 |  | 146 | 44 | 13 |  |
|  |  | 50 - <60 | 73<br>(20.3%) | 19 | 54 |  | 50 | 10 | 13 |  |
|  |  | 60 - <70 | 16 (4.4%) | 8 | 8 |  | 7 | 1 | 8 |  |
|  |  | 70 - <80 | 7 (1.9%) | 2 | 5 |  | 2 | 3 | 2 |  |
|  |  | >80 | 10 (2.8%) | 4 | 6 |  | 6 | 1 | 3 |  |
| 4 | Marital Status | married | 154<br>(42.8%) | 86 | 68 | 0.05 | 115 | 20 | 19 | 0.07 |
|  |  | unmarried | 206<br>(57.2%) | 94 | 112 |  | 135 | 46 | 25 |  |
| 5 | Night Sleep Time | 10 pm or before | 32 (8.9%) | 19 | 13 | <0.001 | 24 | 5 | 3 | 0.02 |
|  |  | 10 pm - 12 am | 238<br>(66.1%) | 101 | 137 |  | 170 | 47 | 21 |  |
|  |  | after 12 am | 90<br>(25.0%) | 60 | 30 |  | 56 | 14 | 20 |  |
| 6 | Waking time | 6 am or before | 87<br>(24.2%) | 40 | 47 | <0.001 | 60 | 14 | 13 | 0.05 |
|  |  | 6-8 am | 230<br>(63.9%) | 107 | 123 |  | 167 | 42 | 21 |  |
|  |  | after 8 am | 43<br>(11.9%) | 33 | 10 |  | 23 | 10 | 10 |  |
| 7 | Night sleep duration (hours) | < 5 | 1 (3%) | 1 | 0 | 0.66 | 0 | 0 | 1 | 0.17 |
|  |  | 5-6 | 125<br>(34.7%) | 68 | 57 |  | 83 | 23 | 19 |  |
|  |  | 6-7 | 183<br>(50.8%) | 88 | 95 |  | 133 | 32 | 18 |  |
|  |  | 7-8 | 40<br>(11.1%) | 18 | 22 |  | 26 | 8 | 6 |  |
|  |  | >8 | 11 (3.1%) | 5 | 6 |  | 8 | 3 | 0 |  |
| 8 | Nap duration (min) | 15 | 20 (5.6%) | 5 | 15 | <0.001 | 14 | 5 | 1 | 0.09 |
|  |  | 15-30 | 80<br>(22.2%) | 40 | 40 |  | 54 | 16 | 10 |  |
|  |  | 30-60 | 119<br>(33%) | 47 | 72 |  | 88 | 13 | 18 |  |
|  |  | 60-90 | 13 (3.6%) | 3 | 10 |  | 9 | 3 | 1 |  |
|  |  | 90-120 | 4 (1.1%) | 1 | 3 |  | 2 | 1 | 1 |  |
|  |  | None | 124<br>(34.4%) | 84 | 40 |  | 83 | 28 | 13 |  |
| 10 | Age (median ± IQR) |  | 26±31 | 27±31 | 25±28 | 0.13 | 27±31 | 25±24 | 26±23 | 0.02 |
| 11 | Day shift frequency (median ± IQR) |  | 20±30 | 20±30 | 16±30 | 0.01 | 20±30 | 20±30 | 15±26 | 0.08 |
|  |  |  |  | public (n) | private (n) | *p* | morning | evening | night | *p* |
| 12 | Night shift frequency (median ± IQR) |  | 10±30 | 10±21 | 10±30 | <0.001 | 10±21 | 10±23 | 10±26 | 0.02 |
| 18 | SQS score (median ± IQR) |  | 41± 91 | 42±81 | 40±67 | 0.08 | 40±85 | 40±49 | 46±70 | 0.30 |
| 19 | MMSE score (median ± IQR) |  | 23± 30 | 23±30 | 23±21 | 0.18 | 23±21 | 24±30 | 20±15 | <0.001 |

IQR, Interquartile range; MMSE, Mini-mental state examination; SQS, sleep quality scale

### 3.2 Association between sleep patterns and cognition

Spearman’s correlation analysis indicates nap duration (*ρ* = 0.12, p = 0.01), and frequency of sleep cycle (*ρ* = −0.24, p < 0.001) to be significantly associated with nurses’ cognition. The value of Spearman’s rho, between nap duration and MMSE shows a moderate positive correlation, indicating that an increase in nap duration leads to an increase in MMSE scores. The value of Spearman’s rho, between frequency of sleep cycle and MMSE shows a strong negative correlation, indicating that an increase in the order of frequency of the sleep cycle leads to a decrease in MMSE scores. The results are summarized in **Table 2**.

**TABLE 2:**
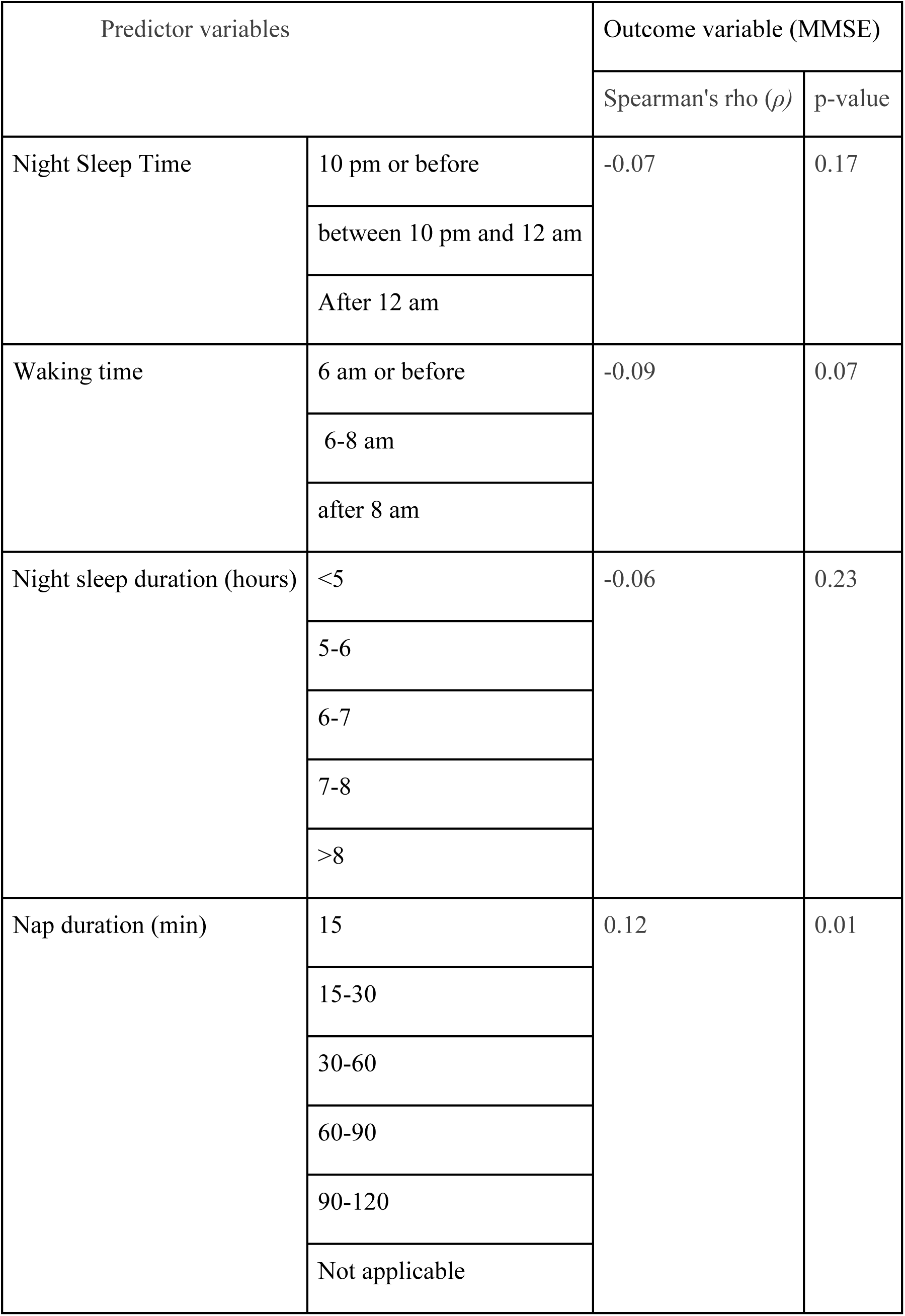
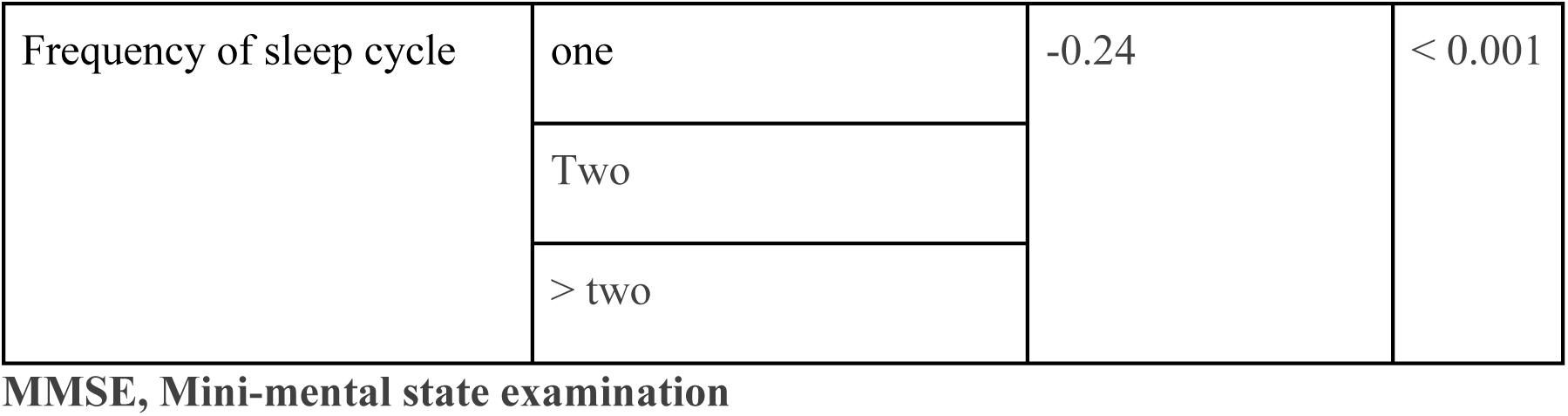
Association between sleep patterns and cognition.

| Predictor variables |  | Outcome variable (MMSE) |  |
| --- | --- | --- | --- |
| | | Spearman's rho ($\rho$) | p-value |
| Night Sleep Time | 10 pm or before | -0.07 | 0.17 |
|  | between 10 pm and 12 am |  |  |
|  | After 12 am |  |  |
| Waking time | 6 am or before | -0.09 | 0.07 |
|  | 6-8 am |  |  |
|  | after 8 am |  |  |
| Night sleep duration (hours) | <5 | -0.06 | 0.23 |
|  | 5-6 |  |  |
|  | 6-7 |  |  |
|  | 7-8 |  |  |
|  | >8 |  |  |
| Nap duration (min) | 15 | 0.12 | 0.01 |
|  | 15-30 |  |  |
|  | 30-60 |  |  |
|  | 60-90 |  |  |
|  | 90-120 |  |  |
|  | Not applicable |  |  |
| Frequency of sleep cycle | one | -0.24 | < 0.001 |
|  | Two |  |  |
|  | > two |  |  |

MMSE, Mini-mental state examination

### 3.3 Association between sleep quality and cognition

Spearman’s correlation test results for the association between sleep quality and cognition indicate a correlation of weak strength between SQS and MMSE, in the negative direction, as explained by the value of Spearman’s rho (*ρ* = −0.024). This shows that an increase in the value of SQS leads to a decrease in MMSE scores. The results are summarized in **Fig. 2**.

**Fig 2:**
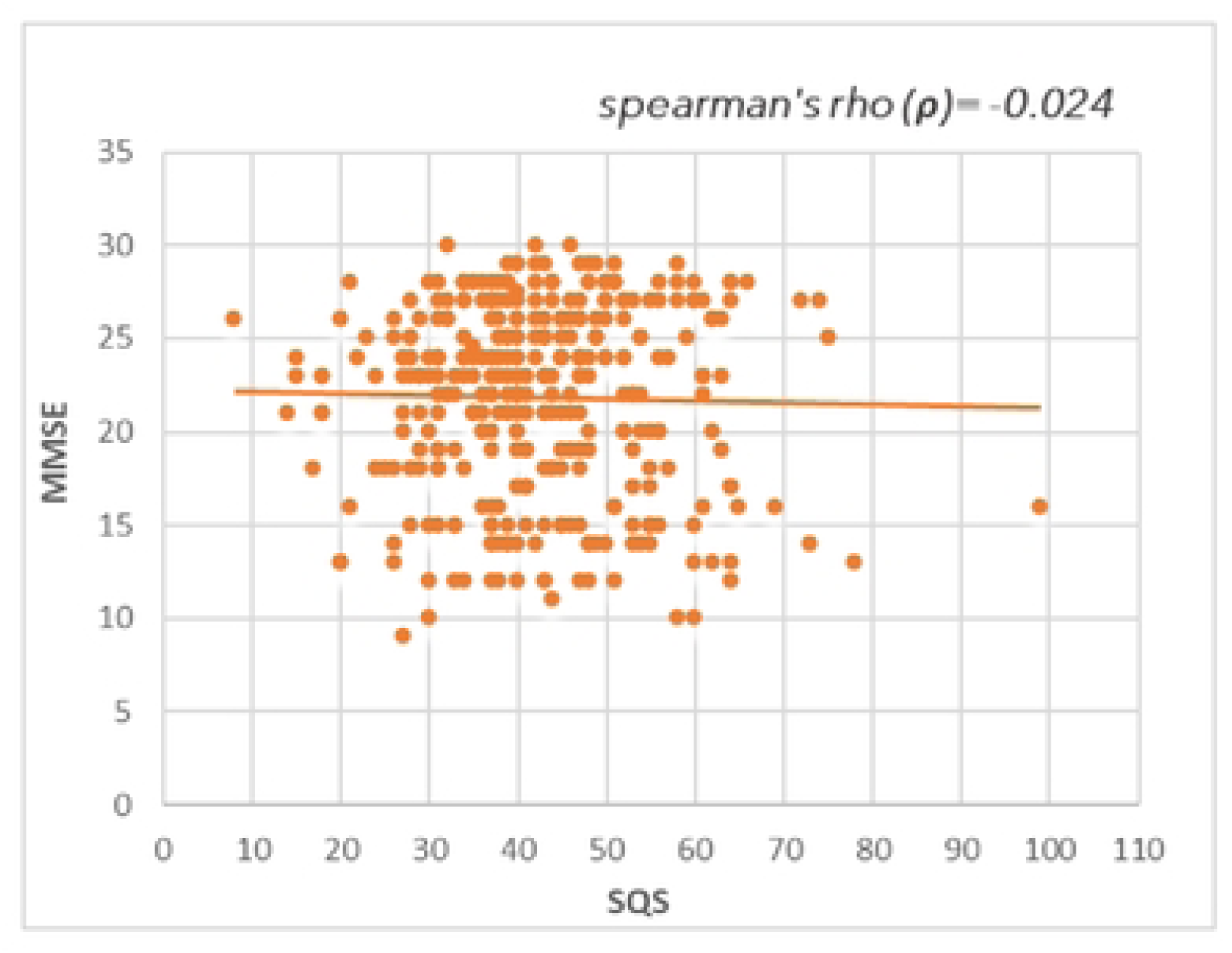
Scatter plot showing the correlation between the sleep quality (SQS) and cognition (MMSE). The spearman’s rho value is mentioned at the top of the figure, showing the magnitude and direction of correlation. MMSE; Mini-mental state examination: SQS; Sleep quality scale

### 3.4 Illustration of association between all the exposures and outcomes

The correlation between all the exposures (demographic and clinical) and the outcomes (hospital type, shift, and cognition) in the study are illustrated in the acyclic graph, present in **Fig. 3**.

**Fig 3:**
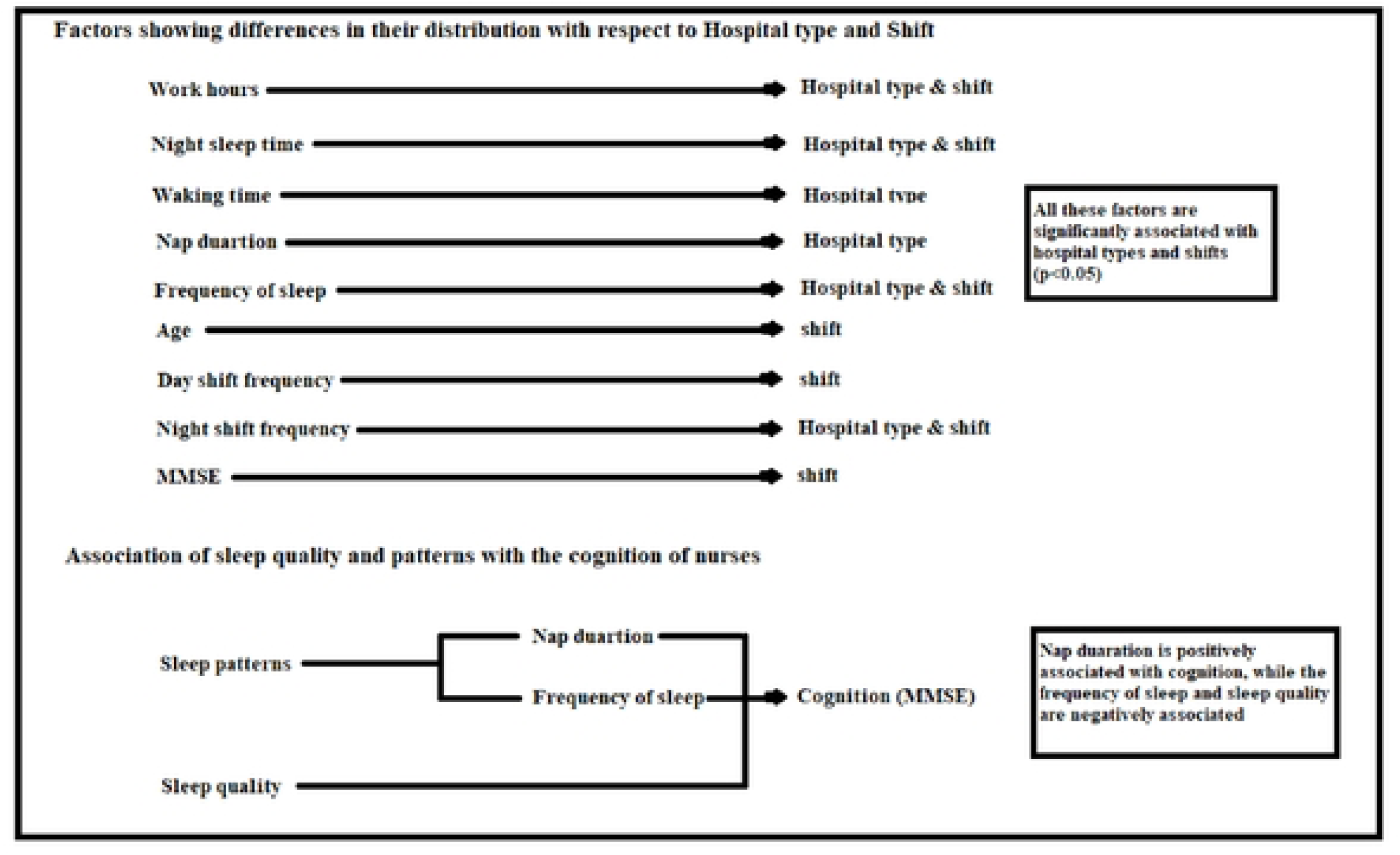
Acyclic graph summarizing the study findings.

## 4.0 Discussion

This study examined the correlation of cognitive abilities of nurses of public and private tertiary care hospitals with their sleep quality and sleep patterns. Sleep and cognition have always been closely tied together, and our results also support these findings. The SQS score for public hospital nurses (42 ± 81) was higher than that of private hospital nurses (40 ± 67), although the p-value was insignificant. Consequently, the MMSE score for public hospital nurses was lower (23 ± 30) compared to the private hospital nurses (23 ± 21). This finding could be related to the higher sleep quality observed among private hospital nurses. A survey conducted on 222,000 subjects revealed that individuals with poor sleep quality (PSQI > 5) exhibited subjective cognitive decline (SCD) approximately twice as often as those with good sleep quality (SCD: Odds ratio [OR] = 1.983, 95% confidence interval [CI] = 1.915-2.054).[22] Moreover, compared to older adults, younger participants demonstrated a greater influence of sleep quality on subjective cognitive decline.

The lower SQS scores indicate that the sleep quality of private hospital nurses was much better. This can be explained by the fact that a dominant portion of the private nurses slept before midnight and woke up before 8 am, compared to public hospital nurses. The p values for both night sleep time (p < 0.001) and wake-up time (p < 0.001) were significant. A study conducted on nurses in Wuhan, China, supports our findings by demonstrating that nurses who sleep before midnight have significantly better overall sleep health [odds ratio (OR): 0.49 (95% CI: 0.29 to 0.80, p = 0.005)].[23] Additionally, a higher number of private hospital nurses slept for an average period of 6-8 hours compared to public hospital nurses for the same duration. A recent study conducted in China concluded that a sleep duration of 6-7 hours was essential for optimized cognitive performance.[24] Another study established an inverted U-shape correlation between the duration of sleep and global cognitive decline. Participants sleeping for ≤ 4 hours (P = .001) and ≥ 10 hours (P = .003) each night had declining cognitive scores compared to the reference group (7 hours per night. [25]

Another factor contributing to the private hospital nurses having superior sleep quality and higher cognitive abilities could be their increased nap duration. The effect of nap duration on the sleep quality of nurses is proven by a study conducted on 196 nurses in Japan. The results show that in comparison to those with a total nap duration of < 90 min, the nurses with a nap duration of ≥ 90 min experienced less drowsiness after nap breaks and decreased fatigue at the end of the shift.[26] The p-value for nap duration between the public and private hospitals was also significant (p= 0.00). A systematic review and meta-analysis conducted in 2023 concluded that daytime naps taken in the range of 30 and < 60 minutes had a moderate-to-high effect on boosting cognitive performance and minimizing perceived fatigue levels.[27] Another systematic review, combining the results of 15 studies conducted on nurses, confirmed that increased daytime napping has a positive effect on cognition.[28]

Yet another factor accounting for private hospital nurses having lower SQS scores and in turn, having better cognition is the day shift (p=0.01) and night shift frequency (p<0.001). The nurses in the private hospitals were averaging a total of 10±30 days of night shifts per month compared to the 10±21 days per month of public hospital nurses. Similar results were found in a study conducted on 222 nurses in different public and private sector hospitals in Karachi. The study stated that the majority of nurses (56.8%) had greater than 8 shifts per month, on average.[29] A study conducted in 2019 on 23 rotating shift nurses revealed that not only did sleep disturbance and sleep-related impairment increase, but the neurobehavioral performance, particularly in the domains of memory, also decreased in all the nurses following night shifts compared to after working day shifts.[30] Similar results were found in a study conducted on physicians working the night shift.[31]

One of the other objectives of our study was to evaluate any notable change in the quality of sleep and cognition across different shift nurses. The p-value for the MMSE score (median ± IQR) was highly significant (p < 0.001) with evening shift nurses having the highest cognition scores (24 ± 30), followed by day shift nurses (23 ± 21), and night shift nurses (20 ± 15). These findings are supported by a survey conducted on 228 nurses in Jinhua Municipal Central Hospital in China, which reported that most night shift nurses worked > 50 hours (56.26 ± 20.25) (mean ± SD), while most day shift nurses worked < 50 hours (41.69 ± 16.66) (mean ± SD).[32] A study conducted in 2021 on shift work strengthens our findings, demonstrating that not only do longer working hours (≥ 55 h/week) negatively impact cognitive abilities, but long night shifts disrupt sleep quality.[33] Another study, conducted on nurses working more than 50 hours per week for consecutive days found decreased cognition levels and increased sleepiness in those nurses (p = 0.00).[34] The p values for both night sleep time and wake-up time amongst the shift nurses were significant (p = 0.02, p = 0.05, respectively). These findings could further explain why evening shift nurses had the lowest average SQS scores and the highest MMSE scores compared to night shift nurses. A study comprising 4,395 school-going children portrayed that participants with an early bed and wake-up led to higher academic performance.[35]

When sleep patterns were compared with cognition, the two main variables that proved significant were frequency of sleep cycles (p < 0.001) and duration of nap (p = 0.01). Our study demonstrated a negative correlation between the sleep cycle frequency and cognition (ρ = −0.24). These findings contradict a study conducted in the US, on 90 nurses, which reported a positive correlation between cognition and amount of sleep of nurses.[36] Our study also found a positive relation between nap duration and cognitive scores (ρ = 0.12), indicating moderate/ longer nap duration was positively correlated with MMSE scores. A study conducted in Korea strengthens our findings, stating that cognitive test scores were higher in nappers than in non-nappers (*p*<0.001). Furthermore, participants with a sleep debt of > 60 minutes benefited more from longer than shorter naps.[37]

Finally, the relation between overall sleep quality, among nurses in public and private tertiary care hospitals, with cognition was also confirmed. The findings indicated a negative correlation, where high SQS score was inversely related to MMSE scores (*ρ* = −0.024). Several studies support these findings. One study emphasized that maintaining good sleep quality in young adulthood and middle age is crucial to improved cognitive functioning and the foremost protector against age-related cognitive decline.[38] Another research conducted on 196 nurses found that 41.8% (95% CI: 41.8-55.6) of participants had poor sleep quality and the prevalence of common mental disorders was identified in 36.7% (95% CI: 30.1-43.9) of them.[39] Yet another study conducted in 2021 in Korea revealed that poor sleep quality was closely related to subjective cognitive decline and the functional limitations associated with it.[40]

Our study has some limitations. First, the cross-sectional design of this study did not allow us to estimate potential long-term associations between the poor sleep quality, sleep patterns, and the cognitive abilities of the nurses. Secondly, as there were no pre-validated tools for the assessment of sleep patterns of the nurses according to our objectives, we used a self-administered questionnaire for the assessment of sleep patterns. However, we conducted a pilot study under proper protocols and made the necessary changes before implementing the questionnaire. We acknowledge these limitations and we are hoping that future researchers who are interested in this topic cover the gaps in our study.

## 5.0 Conclusion

The results of this study conclude that sleep plays an important role in the cognitive functioning of nurses in tertiary care hospitals. There is a strong positive correlation between the duration of naps taken and cognition. The results show a significant difference in the cognitive performance of shift nurses, with evening shift nurses demonstrating the best cognition and sleep quality, while night shift nurses had the worst cognition and sleep quality. Changes must be made to the work schedule of nurses employed in tertiary care hospitals to increase their sleep quality and, in effect, cognition, for the safety and well-being of both healthcare professional and patients.

## 6.0 Statements & Declarations

### 6.1 CRediT AUTHOR STATEMENT

Anusha Azhar: visualization, writing-original draft, methodology Muhammad Maaz bin Zahid: conceptualization, validation, formal analysis Sunnia Shah: investigation, writing-review and editing Rida Islam: Writing, review and editing, Sakhshi Kumari: writing-original draft, investigation Muhammad Sanaan Noor: Data curation and analysis Aina Khushbakht Tariq: Data curation and editing Bushra Iftikhar: supervision and project administration

The corresponding author (Muhammad Maaz bin Zahid) is also responsible for all content as a guarantor.

### 6.2 Funding

This research received no specific grant from any funding agency in the public, commercial, or not-for-profit sectors.

### 6.3 Competing Interests

All the authors have no competing interests to disclose.

### 6.4 Checklist

STROBE checklist was followed during our study.

### 6.5 Data availability statement

The data that has been used is confidential. The research respondents were assured raw data would remain confidential and would not be shared.

All relevant data to the study can be accessed through contact with the corresponding author.

### 6.6 Ethics statement

- **Patients’ consent:** The patient’s consent for publication was not required.
- **Ethics approval:** Ethical approval was obtained from the Institutional Research and Ethical Review Board (IREB) of Khyber Medical College (KMC) on 13th February 2024 (No. 87/DME/KMC). Study details were explained, and informed consent was obtained from guardians/parents before the initiation of the study.

## Notes

### Competing Interest Statement

The authors have declared no competing interest.

### Author Declarations

Ethical approval was obtained from the Institutional Research and Ethical Review Board (IREB) of Khyber Medical College (KMC) on 13th February 2024 (No. 87/DME/KMC). Study details were explained, and informed consent was obtained from guardians/parents before the initiation of the study.

